# *H*emorrhage *E*valuation And *D*etector *S*ystem for *U*nderserved *P*opulations: *HEADS UP*

**DOI:** 10.1101/2023.06.25.23291870

**Authors:** Saif Salman, Qiangqiang Gu, Benoit Dherin, Sanjana Reddy, Patrick Vanderboom, Rohan Sharma, Lin Lancaster, Rabih Tawk, William David Freeman

## Abstract

**Introduction:** Intracerebral hemorrhage (ICH) is the second most common cause of stroke and remains the second leading cause of disability impacting underserved areas. Since 2015, there has been a paradigm shift in managing ischemic stroke through applying AI and ML. However, ICH patients lack such protocol.

**Objective:** To create a rapid, cloud-based, and deployable ML method to detect ICH potentially across the Mayo Clinic enterprise then expand to involve underserved areas.

**Methods:** We utilized RSNA dataset for ICH. We made four total iterations using Google Cloud Vertex AutoML. We trained an AutoML model with 2,000 images followed by 6,000 images from both ICH positive and negative classes. Pixel values were measured by the Hounsfield units presenting a width of 80 Hounsfield and a level of 40 Hounsfield as the bone window. This was followed by a more detailed image preprocessing approach by combining the pixel values from each of the brain, subdural, and soft tissue window-based grayscale images into R(red)G(green)B(blue)-channel images to boost the binary ICH classification performance. Four experiments with AutoML were applied to study the impacts of training sample size and image preprocessing on model performance.

**Results:** Out of the four AutoML experiments, the best-performing model achieved a 95.8% average precision, 91.4% precision, and 91.4% recall. Based on this analysis, our binary ICH classifier ***HEADS UP*** is both accurate and performant.

**Conclusion:** ***HEADS UP***, is a rapid, cloud-based, deployable ML method to detect ICH. This tool can help expedite the care of patients with ICH in resource-limited hospitals.

## Introduction

Strokes are the second most common cause of death and disability globally ^1^. Despite the ischemic subtype being more prevalent (80-85%), the intracerebral hemorrhage (ICH) subtype is considered deadlier and more disabling. ICH carries an average mortality rate of 40% by 30 days ^2^ and can reach up to 60% in one year ^3^. Increasing age, female gender, rurality, and history of chronic illnesses are risk factors for increased ICH mortality ^4^.

Primary ICH can be caused by hypertension, atherosclerosis, or amyloid angiopathies. To a lesser extent, ruptured aneurysms, bleeding tumors, vascular malformations, coagulopathies, and previous thrombolysis can cause secondary ICH ^5^. Patients develop a myriad of short and long-term complications ranging from re-bleeding, expansion, vasospasm, seizures, cognitive decline, and multi-systemic neurological injuries inflicted on the myocardium and lungs ^5^.

Early detection directing timely intervention is paramount and crucial in determining outcome ^6^. Regrettably, while medical history can be the first guide towards unraveling the etiology of strokes ^7^, sole clinical examination lacks a particular role in distinguishing ischemic from hemorrhagic strokes as the presenting symptoms can be somewhat similar. Hence, non-contrast head CT scans (NCCT) remain the gold standard differentiation method ^5,8–11^.

Machine learning (ML) utilizes unique models such as feedforward artificial neural networks (ffANNs), Random forest (RF), support vector machine (SVM), logistic regression (LR), Stacked Convolutional Denoising Auto-encoders, Principal Component Analysis (PCA), and Multi-layer Perceptron (MLP). The aforesaid feedforward artificial neural networks (ffANNs) is the most utilized to predict case-specific outcomes competing with the decisions made by clinicians ^12–14^.

Since 2018, there has been a paradigmatic change in the management of ischemic strokes through the implementation of Artificial Intelligence (AI) and ML methods in processing NCCT and CT perfusion scans to predict large vessel occlusion (LVO) in the prehospital setting, improve precise detection of clots, decrease latency until intervention, improve reperfusion and predict clinical outcomes ^15–20^.

ICH patients lack such a predictive model in a streamlined system to improve the acute stroke systems of care, earlier detection, and outcome. There is an unfulfilled need to create an earlier ICH detection method that accelerates interventions. <u>H</u>emorrhage <u>E</u>valuation <u>A</u>nd <u>D</u>etector <u>S</u>ystem for <u>U</u>nderserved <u>P</u>opulations (***HEADS UP)*** is a high-precision, rapid, and cloud-based deployable model created using Google Cloud VertexAI AutoML to detect ICH remotely. Hence, ***HEADS UP*** can detect ICH in a time-efficient manner, makes the most of our health system, and improves outcomes. We aim to implement ***HEADS UP*** across the Mayo Clinic enterprise and expand to involve underserved areas.

The main novelty in ***HEADS UP*** is based on the incorporated joint efforts between ML scientists, neurologists, and neurosurgeons to generate a deployable and high-precision model within two weeks and individualize towards patient care, future discovery, and translation of sciences.

## Materials and methods

### Dataset

We aim to develop a binary image classifier to identify positive ICH cases in two-dimensional NCCT images. We downloaded the original dataset from the Radiological Society of North America (RSNA) intracranial hemorrhage database ^21^. The images in the dataset were categorized into seven annotation groups representing the five known intracranial hemorrhage subtypes (intraparenchymal, subarachnoid, subdural, intraventricular, and epidural), along with another two groups that include any type of hemorrhage, and none hemorrhage ^21^. Totally, we attained 752,803 labeled images (**Table 1**).

**Table 1.**
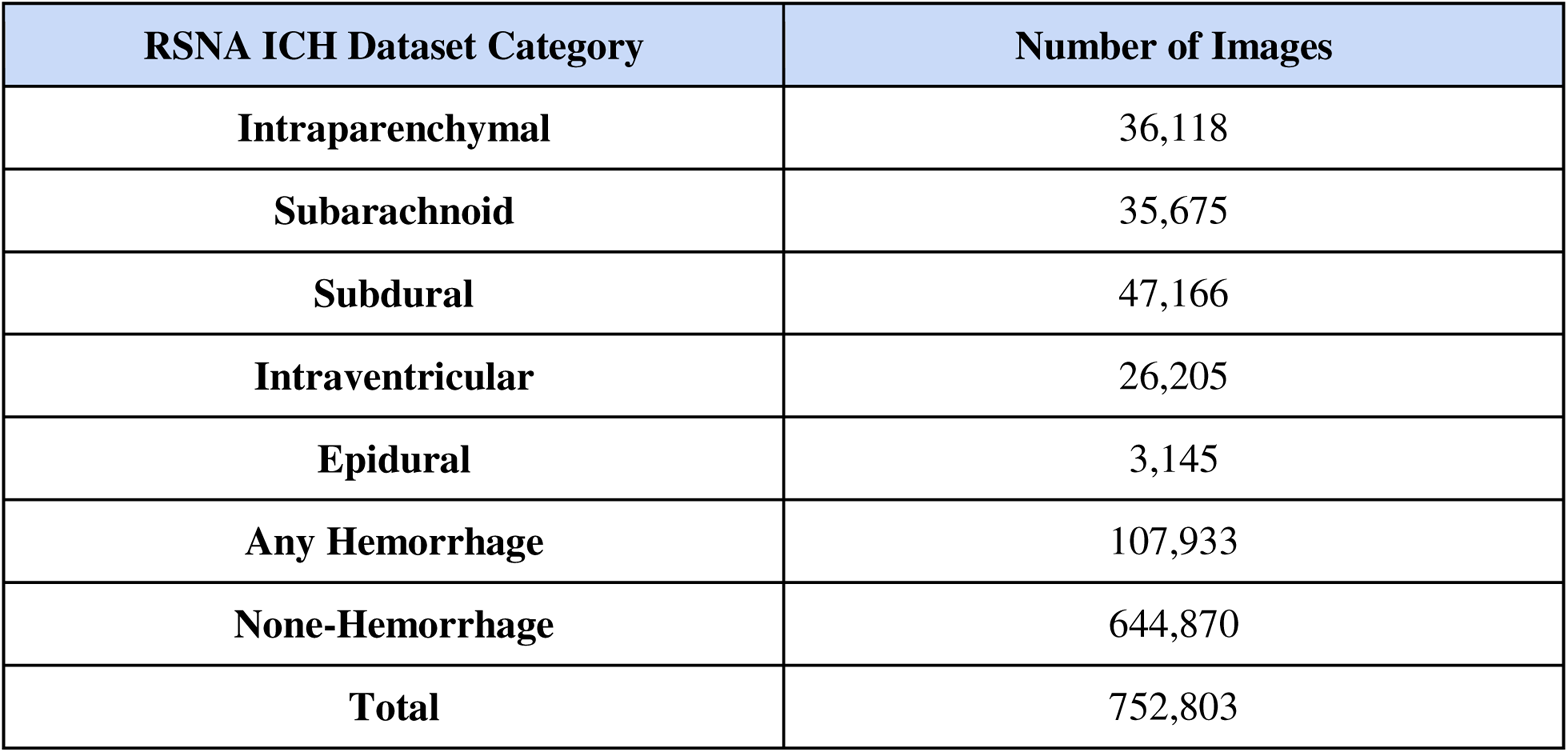
Summary of RSNA Intracranial Hemorrhage Dataset Information.

| RSNA ICH Dataset Category | Number of Images |
| --- | --- |
| Intraparenchymal | 36,118 |
| Subarachnoid | 35,675 |
| Subdural | 47,166 |
| Intraventricular | 26,205 |
| Epidural | 3,145 |
| Any Hemorrhage | 107,933 |
| None-Hemorrhage | 644,870 |
| Total | 752,803 |

### Data Pre-Processing

After excluding duplicates, we pooled the images from the six hemorrhage subtypes as a single ICH positive class and images from the non-hemorrhage group as the ICH negative class to apply a binary image classifier and detect ICH cases when present. Two thousand randomly selected images were used for model development and testing as one thousand images from each ICH positive and negative class. To better understand the impact of training sample size on model performance, we created another dataset using the same method but with 6,000 images, including 3,000 images from each ICH positive and negative class.

Original image data from the RSNA hemorrhage dataset was saved as Digital Imaging and Communications in Medicine (DICOM) standard format ^22^. Since the VertexAI AutoML did not directly support the DICOM format, the raw image pixel arrays were extracted from the DICOM files and were saved as PNG formats.

We further arranged each image into brain, subdural, and soft tissue windows with different Hounsfield units. The difference in the Hounsfield units enhanced further differentiation between ICH and non-ICH cases. An additional approach combined the pixel values from each of the brain, subdural, and soft tissue window gray-scale images into three-channeled R(red)G(green)B(blue) images.

To be more precise, each of the original PNG format pixel arrays was filtered into brain, subdural, and soft tissue windows based on their unique window width and level values in Hounsfield units. The brain window has a width value of 80 Hounsfield units and a level value of 40 Hounsfield units. The subdural window has a width value between 130 and 300 Hounsfield units and a level value between 50 and 100 Hounsfield units. The soft tissue window has a width value of 250 to 400 Hounsfield units and a level value of 50 Hounsfield units ^23,24^. Then, the 512 x 512-dimensional pixel arrays of the brain, subdural, and soft tissue window gray-scale images were placed into the red, green, and blue channels to form the RGB images.

The final combined three-channeled RGB images were used to train AutoML-based binary ICH classifiers. The images with the corresponding labels were randomly split into 90% for model development and 10% for model testing and applied to each patient level.

### Model Training and Testing

Images within the model development sets were used to optimize model predictions during the training phase. However, images in the testing set were used to evaluate the performance of the developed model.

We applied the XRAI method, the latest approach for AI model interpretation. XRAI utilizes region-based image attribution to determine regions from the inference images with the highest predictions for binary ICH classes ^25^. Further, XRAI combines the integrated gradients method with additional steps to determine regions with the highest contributions for class prediction. The implementation of this study was programmed in Python (Python 3.11), and the data pre-processing source code is publicly available at https://github.com/quincy-125/MG-HEADSUP.

Four experiments were executed to fully understand the impacts of training sample size and data pre-processing on AutoML-based binary ICH classification performance (**Supplementary Table 1**). We started by increasing the training sample size from 2,000 to 6,000. We further trained the binary ICH classifier on gray-scale images with pixel values filtered by the Hounsfield unit range corresponding to the brain window. This was followed by training the binary ICH classifier on the combined three-channeled RGB images.

The first experiment was training the AutoML model on 2,000 gray-scale brain window images (E1). The following three experiments used the combined three-channeled RGB images to train the binary ICH classifier. The second experiment (E2) assigned equal weight to every pixel from the red, green, and blue channels. The resulting pixel values from the combined three-channeled RGB images were saved as floating numbers with 64 bits (*float64*). The third experiment (E3) is similar, but the resulting pixel values from the combined three-channeled RGB image were saved in the format as integers with eight bits (*uint8*).

The last experiment (E4) performed best by duplicating the pixels from each brain, subdural, and soft tissue window gray-scale image to generate window-based combined RGB images with heavily-weighted red channel pixel values. We further combined each of the three window-based RGB images and produced the final combined equally-weighted RGB image with each pixel value saved in the *float64* format (**Figure 1**).

**Figure 1.**
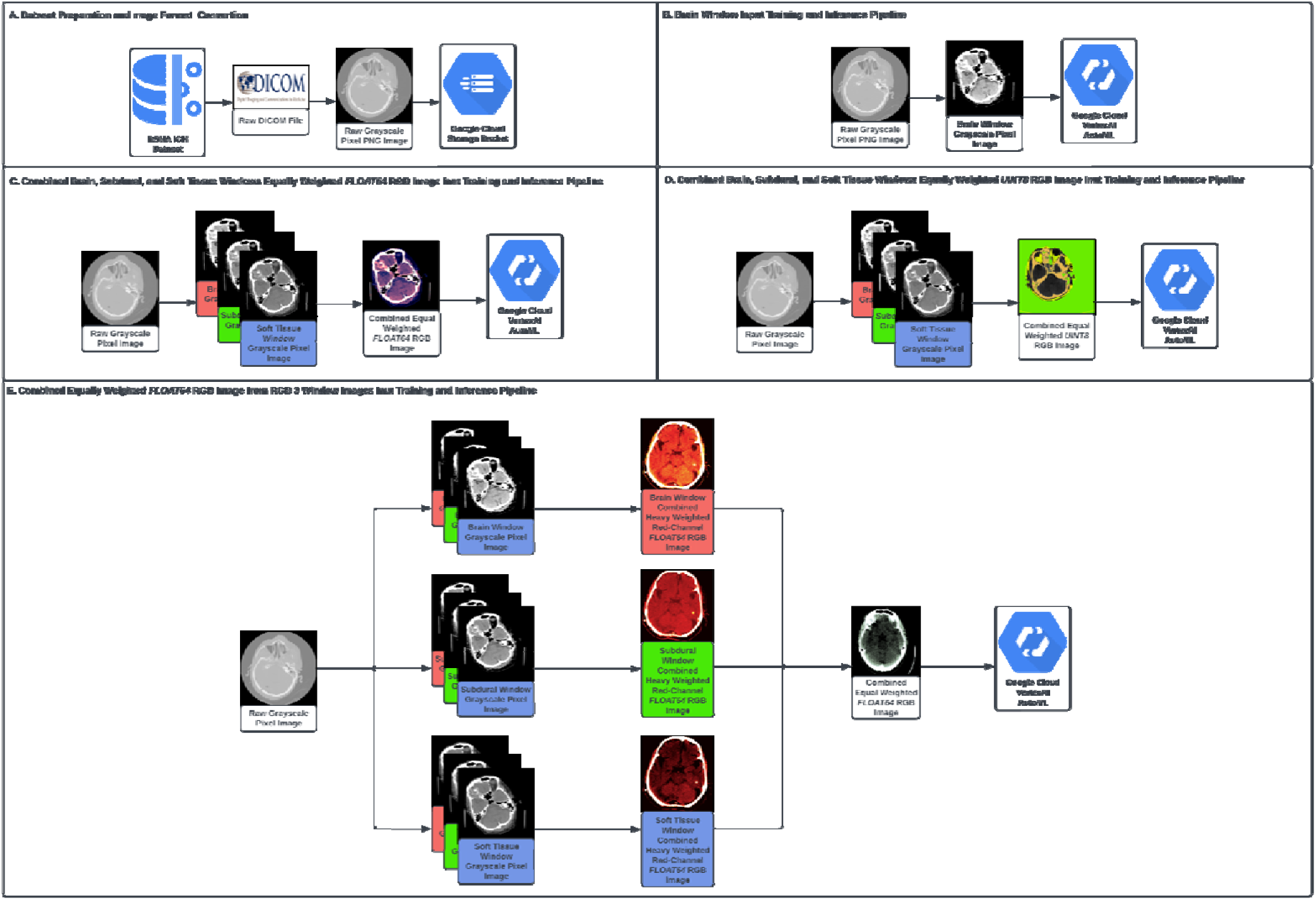
Diagram of the entire dataset preparation, image pre-processing, model training and inference pipeline. A). Dataset preparation including downloading original DICOM format files from the RSNA dataset, and converting DICOM format image into PNG format; B). Image pre-processing for AutoML training and inference pipeline, which corresponds to the experiment indexed as E1 in **Supplementary Table 1**; C). Image pre-processing for AutoML training and inference pipeline, which corresponds to the experiment indexed as E2 in **Supplementary Table 1**; D). Image pre-processing for AutoML training and inference pipeline, which corresponds to the experiment indexed as E3 in **Supplementary Table 1**; E). Image pre-processing for AutoML training and inference pipeline, which corresponds to the experiment indexed as E4 in **Supplementary Table 1**.

## Results

Precision and recall values were reported for each of the four experiments discussed in the **Model Training and Testing Section** with a threshold value of 0.5. In addition, average precision (the area under the precision-recall curve) was considered as a measurement of our models’ performance. All of the reported statistical metrics regarding each of the four experiments are listed in **Table 2**.

**Table 2.** Inference results for each of the four experiments.

| Experiment Index | Recall<br>(Threshold=0.5) | Precision<br>(Threshold=0.5) | Average Precision |
| --- | --- | --- | --- |
| E1 | 88.90% | 88.90% | 91.50% |
| E2 | 90.00% | 90.00% | 95.20% |
| E3 | 90.00% | 90.00% | 95.20% |
| E4 | **91.40%** | **91.40%** | **95.80%** |

## Discussion

ICH is a medical emergency that requires vigorous intervention, particularly with hemorrhage in deep structures such as the basal ganglia, pons, or cerebellum. Elderly patients are more prone to amyloid angiopathy, causing lobar hemorrhages ^26^. Despite the etiologies, the use of anticoagulants, especially if combined with the aforementioned, potentiates the risk of expansion and rebleeding with a 0.6% risk or less per year ^27,28^.

Headache as a presenting sign is followed by loss of function that can be seen in both ischemic and hemorrhagic strokes, despite being more common in the latter. Hematoma expansion within the confined intracranial space drastically increases the intracranial pressure (ICP), jeopardizing blood flow and culminating in herniation. This mass effect is reflexively accommodated by the increase in mean arterial pressure (MAP) to overcome the increased ICP and ensure cerebral perfusion. However, Increased MAP can contribute to hematoma expansion, and it may ultimately fail to ensure perfusion due to an exponential increase in ICP with worsening hemorrhage. Hence, urgent interventions to decrease ICP and expansion of the hematoma, such as mannitol or hypertonic saline infusion, raising the head of the bed, and optimal blood pressure control, are mandated to prevent further herniation, loss of function, and death ^5,27,28^.

Cerebral edema is a fatal complication that may last for weeks. It begins with a clot contraction phase in the first few hours with mottled appearance on NCCT scans. Upon further coagulation, it peaks in the next 48 hours with hyperdensity. As ICH is resorbed, NCCT scans will show the hypodensity of the lesion. However, continuous expansion may render secondary involvement of the ventricles, a potentially fatal complication with a high 30-day mortality rate of 40% that may result from hydrocephalus unless diagnosed early and intervened timely ^5,29,31^.

ICH causes the highest stroke mortality rates in rural areas and underserved populations. This can be attributed to limited health education, primary prevention, and access to healthcare. However, the need for early detection due to absent transportation methods, imaging facilities, or healthcare professionals capable of interpreting the imaging results is still the most profound factor affecting management and outcome ^31–37^. While modifiable risk factors remain a focus of interest, providing a streamlined approach to expedite ICH diagnosis followed by guided management in the acute phase is critical to improve outcomes.

Given the rising era of AI and ML in various medical fields and the application of ML in diagnosing ischemic strokes and predicting LVO, we perceive similar roles in processing NCCT scans and diagnosing ICH. These roles improved accuracy, time latency, and outcome. Davis et al. incorporated an ML algorithm, “Aidoc,” based on a convolutional neural network (CNN) to detect ICH on the NCCT scans. They reported reduced turnaround time to detect hemorrhage during the acute phase, while no change in the length of emergency department stay compared to non-ICH patients ^38^. Arbabshirani et al. aimed to develop a model capable of detecting ICH through large numbers of head NCCT scans and predict the reduction in time taken for interpretation. They reported decreased latency for interpretation by 96% and the ability to identify missed hemorrhage by radiologists ^39^. Further, Lyu et al. used logistic regression analysis of NCCT scans for 238 patients to report a diagnostic accuracy that surpasses radiologists when differentiating between primary and secondary ICH. They also reported that lobar hemorrhages in females could be correlated with secondary ICH, while hypertension will most likely cause primary ICH in all patients ^40^.

We created ***HEADS UP*** using Google Cloud VertexAI AutoML, which automatically identifies the most appropriate models to maximize the binary ICH classification performance. It is a rapid, cloud-based, and deployable ML method to detect ICH. The co-author (WDF) used BARD, a conversational generative artificial intelligence chatbot developed by Google, to generate the acronym ***HEADS UP***. Hence, drawing attention to the currently limited care in underserved areas around the globe regarding ICH, a debilitating disease requiring timely intervention and efficient management (**Figure 3**).

Out of the four experiments conducted through our AutoML model, experiment E4 was conducted on 6,000 combined RGB images and achieved the highest values regarding average precision (95.80%), optimal precision (91.40%), and recall (91.40%). This emphasizes the significance of levitating the capability of AutoML in detecting ICH-positive cases from NCCT images. In addition, experiment E4 outperformed the remaining three experiments with a 4.30%, 0.60%, and 0.60% boost in average precision. This suggests that increasing the training sample size with appropriate image preprocessing could boost the performance of binary ICH classification.

However, given the ability of XRAI to interpret AutoML predictions, we believe that the annotations provided by the RSNA dataset were not entirely accurate. For instance, in **Figure 2**, one of the testing results using XRAI in E3 considered the available image extracted from the RSNA dataset as ICH negative. At the same time, our AutoML model predicted it as ICH positive and labeled it with a yellow polygon. This was consistent with a subsequent evaluation by a senior neurosurgeon in our department, who used the red polygon shown in **Figure 1** to highlight the same lesion. Further, this red polygon contained the yellow polygon that was already highlighted by the XRAI and was confirmed as a true positive ICH lesion. We believe this false positive prediction is due to the incorrect labeling in the RSNA dataset.

**Figure 2.**
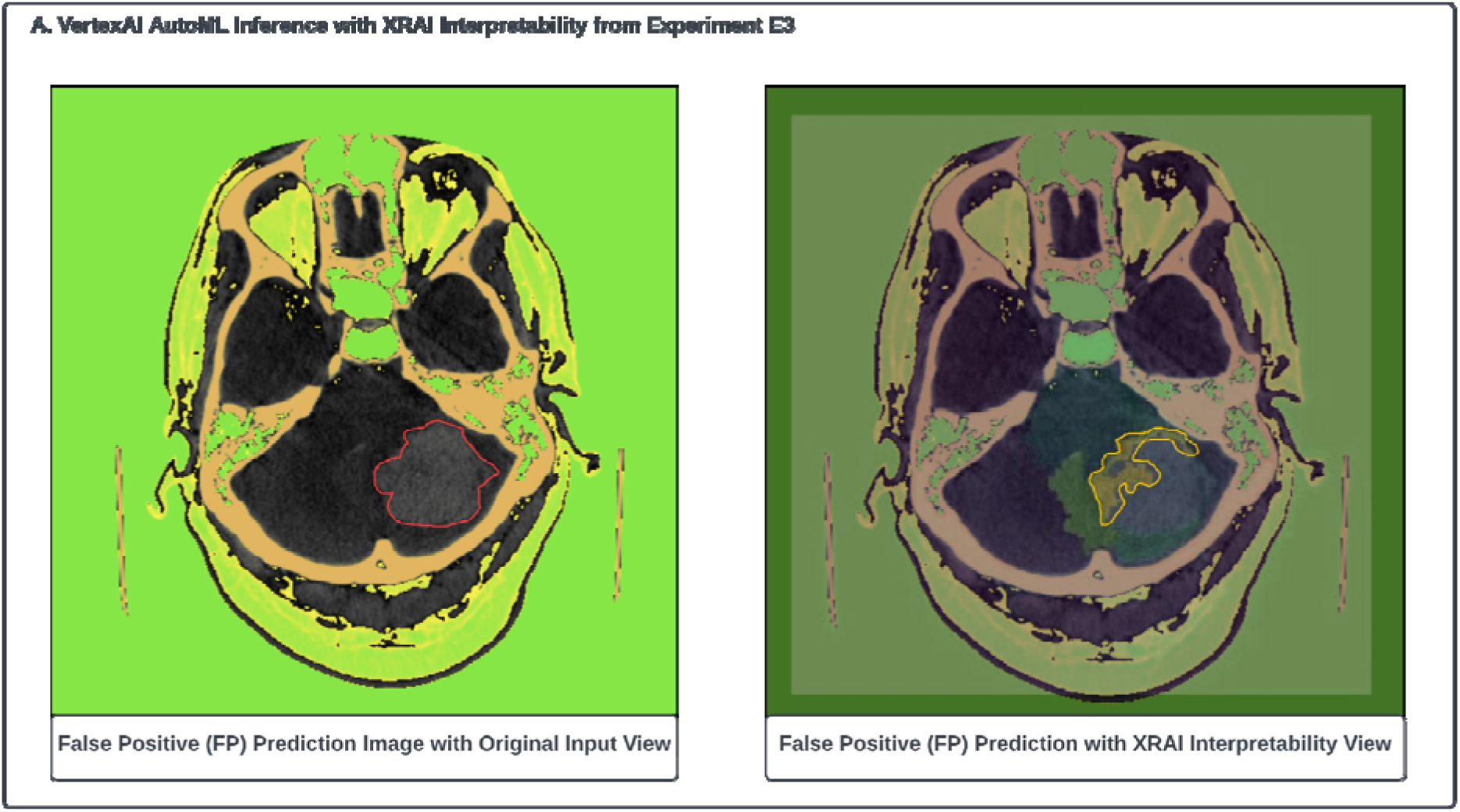
VertexAI AutoML inference results from the experiment E3. A). One false positive prediction inference example derived from experiment E3 with the XRAI interpretation results. The red polygon was the annotation provided by a senior neurosurgeon at the Mayo Clinic with the true ICH positive lesion. The yellow polygon was highlighted by the XRAI corresponding to the false positive prediction from the VertexAI AutoML model.

**Figure 3:**
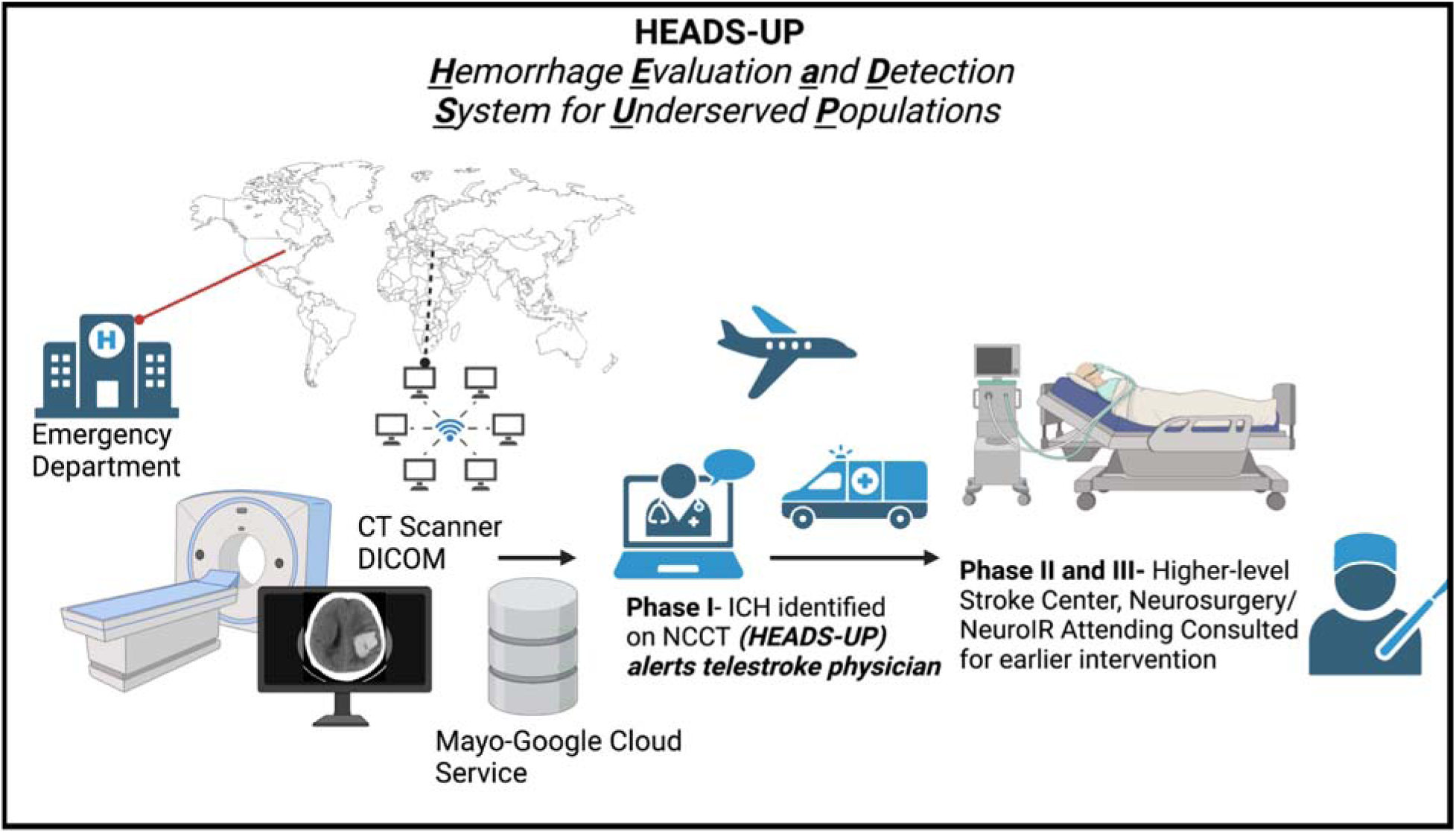
Courtesy of Co-Author WDF, describes the role of ***HEADS UP*** to detect ICH in the underserved areas and expedite efficient and timely intervention.

The incorrect labels from the RSNA dataset could be the reasons preventing our AutoML model from achieving higher performance metrics. Hence, training an AutoML model with more accurate labels may boost performance. Further, a thorough evaluation is highly recommended for the models pre-trained on the RSNA dataset before clinical implementation.

To our knowledge, this is the first binary ICH classifier that utilized XRAI for model interpretation. XRAI in Google VertexAI AutoML is considered a low-code software development approach, ensuring better team communication despite different backgrounds. Hence, we were able to develop ***HEADS UP*** within two weeks.

Wang et al. reported a similar binary ICH classifier that utilized class activation mapping (CAM) ^41^ to find the regions contributing to predicting the classification model. They directly used 674,258 two-dimension scans for model development and generated a 95.8% recall from a completely different test set than ours ^42^. In comparison, we started applying our model to a small sample size and expanded in size to understand the correlation between sample size and performance more thoroughly. Further, as described previously, we applied XRAI interpretation and manual qualitative evaluation by an expert neurosurgeon. Hence, we concluded that the labels provided in the original RSNA hemorrhage dataset lack complete accuracy. We believe such mislabeling could cause a significant risk of misleading model predictions, especially when directly deploying models trained on the RSNA dataset clinically without a thorough clinical evaluation. Further, we did four experiments (**Figure 1**) that elevated performance metrics with a more complicated image preprocessing, while Wang et al. carried an image preprocessing similar to our E2 and E3 experiments.

Through ***HEADS UP***, we aim to establish independence from third-party vendors by relying on internal expertise and talents, staying ahead of developing technologies, and developing a self-sufficient system to replace external offers. While planning this gradual transition, it is critical to ensure compatibility with general medical knowledge as we gain experience. We further aim to maintain medical evaluation on a case-to-case basis, decreasing our dependency. Hence, top-quality services can be provided directly to underserved areas in need.

## Conclusion

***<u>H</u>***emorrhage ***<u>E</u>***valuation and ***<u>D</u>***etector ***<u>S</u>***ystem for ***<u>U</u>***nderserved ***<u>P</u>***opulations, ***HEADS UP***, is a rapid, cloud-based, and deployable ML method to detect ICH. We are considering implementing this system across the Mayo Clinic enterprise and expanding to involve underserved areas to help patients and their clinical teams while creating scientific and intellectual independence from third-party vendors. Utilizing Google VertexAI AutoML catalyzed the communication between our team members to generate the ***Heads UP*** model within two weeks.

## Data Availability

All data produced in the present study are available upon reasonable request to the authors

## Appendix A. Supplementary Table

**Supplementary Table 1.** Details of the four executed training experiments along with the corresponding training sample size and data pre-processing approaches.

| Experiment Index | Training Sample Size | Image Pre-Processing Approach |
| --- | --- | --- |
| E1 | 2,000 | Grayscale Brain Window Image |
| E2 | 2,000 | Combined RGB-Channel Image with Equal Weighted RGB Channel Pixel Values in *Float64* |
| E3 | 2,000 | Combined RGB-Channel Image with Equal Weighted RGB Channel Pixel Values in *Uint8* |
| E4 | 6,000 | Combined RGB-Channel Image Resulting from Each of the Three Window-based RGB images with a Higher Weighted Red Channel Pixel Values in *Float64* |

